# Glucocorticoids inhibit type I IFN beta signaling and the upregulation of CD73 in human lung

**DOI:** 10.1101/2020.04.01.20049700

**Authors:** Juho Jalkanen, Ville Pettilä, Matti Karvonen, Teppo Huttunen, Jami Mandelin, Markku Jalkanen, Markus Malmberg, Kati Elima, Geoff Bellingan, V. Marco Ranieri, Maija Hollmen, Sirpa Jalkanen

**Affiliations:** Faron Pharmaceuticals Ltd., Turku, Finland; Department of Anesthesiology, Intensive Care and Pain Medicine, University of Helsinki and Helsinki University Hospital, Helsinki, Finland; Pharma Ltd., Turku, Finland; Heart Center, Turku University Hospital and University of Turku, Turku, Finland; Medicity Research Laboratory, Department of Microbiology and Immunology, University of Turku, Turku, Finland; Critical Care, University College London Hospitals, NHS Foundation Trust and NIHR Biomedical Research Centre at University College London Hospitals NHS Foundation Trust and University College London, London, UK; Alma Mater Studiorum – Università di Bologna, Dipartimento di Scienze Mediche e Chirurgiche, Anesthesia and Intensive Care Medicine, Policlinico di Sant’Orsola, Bologna, Italy; Institute of Biomedicine, University of Turku, Turku, Finland

**Author notes:** Corresponding author: Prof Sirpa Jalkanen, Medicity Research Laboratory, Department of Microbiology and Immunology, University of Turku, Turku, Finland. equal contribution (Juho Jalkanen, Ville Pettilä, Maija Hollmen, Sirpa Jalkanen).

**Keywords:** CD73, endothelium, ARDS, interferon, glucocorticoids

## Abstract

**Purpose:** Glucocorticoids are widely used to treat acute respiratory distress syndrome (ARDS) despite its use is highly controversial based on randomized controlled trials and meta-analyses. As type I interferons (IFNs) are our first line of defense against severe viral respiratory infections, we explored whether glucocorticoids interfere with IFN signaling and whether their use associates to outcome of IFN beta treatment of ARDS.

**Methods:** We performed a propensity-matched post-hoc-analysis using data from the recent randomized INTEREST-trial comparing IFN beta-1a to placebo in ARDS patients. Based on the results of these analyses we utilized human lung tissue and human pulmonary endothelial cell cultures to investigate the effect of hydrocortisone on IFN nuclear signaling and the protein transcription of CD73, a molecule responsible for vascular integrity.

**Results:** We found that hydrocortisone reduces the production, and prevents the nuclear translocation of IRF9, that is required for IFN beta-dependent signaling of multiple IFN-induced genes. In addition, hydrocortisone inhibits IFN beta-dependent upregulation of CD73 in human lung tissue. Additionally, we found that use of glucocorticoids with IFN beta-1a was independently associated with increased mortality (OR 5.4, 95% CI 2.1–13.9, P< 0.001) in the INTEREST-trial.

**Conclusions:** Glucocorticoids inhibit type I IFN beta signaling and the upregulation of CD73 in human lung. This provides the mechanistic basis for the harmful association of glucocorticoids in IFN beta treated patients in the INTEREST-trial. Most importantly, it strongly speaks against the use of glucocorticoids in viral-induced ARDS such as in the current corona virus pandemia.

**Take home message:** Glucocorticoids inhibit type I interferon beta signaling and the upregulation of CD73 that is a key molecule preventing vascular leakage and harmful leukocyte infiltration into the lungs. This work provides the mechanistic basis for the need to avoid glucocorticoids in viral-induced ARDS, in which endogenous interferon is needed to combat the infection and its consequences.

## Introduction

Acute respiratory distress syndrome (ARDS) is a condition characterized by pulmonary inflammation, diffuse pulmonary edema and refractory hypoxemia that may complicate pneumonia, sepsis, trauma and lead to multi-organ failure. ARDS mortality is approximately 35-40% depending on the severity of the disorder [1]. The treatment of ARDS still relies on the management of the underlying disease and supportive care since pharmacological interventions targeting key pathophysiological determinants of ARDS are still not available [2]. The key pathophysiological event of ARDS is an uncontrolled inflammatory response resulting in injury of the endothelial-alveolar barrier with increased pulmonary vascular leakage [1, 3].

Endogenous production of type I IFNs (IFN beta and alpha) is of outmost importance to fight against viral and bacterial infections [4], since it upregulates cluster of differentiation 73 (CD73) expressed on endothelium cells. CD73 is an enzyme controlling endothelial barrier function and leucocyte recruitment to sites of inflammation via the production of adenosine [5-7], a highly anti-inflammatory substance having cardioprotective, neuroprotective, vasodilatory and angiogenic properties [8]. A phase II trial showed that upregulation of CD73 via intravenous administration of recombinant human interferon is associated with a reduction in 28-day mortality in ARDS [9]. A following placebo-controlled phase III trial (INTEREST) showed that intravenous recombinant human interferon beta-1a did not improve outcome in patients with ARDS [10]. However, the study showed that use of corticosteroids at baseline was higher than 50% and post-hoc analyses revealed a statistically significant interaction between baseline corticosteroids and the treatment group [10]. This present study was set out to test the hypothesis that the lack of efficacy of recombinant human IFN beta-1a in patients with ARDS could be related to the interference of glucocorticoids with IFN beta-1a administration and prevent upregulation of CD73. Accordingly, we first performed a post-hoc propensity-matched analysis of the INTEREST trial (ClincalTrials.gov Identifier NCT02622724), and second we investigated the effects of glucocorticoids on IFN signaling and the up-regulation of CD73 expression using human lung organ cultures and human pulmonary endothelial cells *ex vivo*. Although there is preclinical evidence that steroids block endogenous IFN signaling, this work puts this into context for the critical care community treating ARDS.

## Methods

### Post-hoc propensity-matched analyses from the INTEREST trial

The INTEREST trial was a multicenter, randomized, double-blind, parallel-group trial conducted at 74 intensive care units in 8 European countries (from December 2015 to December 2017) that included 301 adults with moderate to severe ARDS. Patients were randomized to receive an intravenous injection of 10 µg IFN beta-1a (144 patients) or placebo (152 patients)[10]. In this analysis we explored the possible interaction between glucocorticoids and IFN beta-1a. We present the Kaplan-Meyer survival curves until day 90 for IFN beta-1a treated patients with and without concomitant (D0-D6) glucocorticoids.

In addition, we performed propensity score (representing the risk to receive glucocorticoids at randomization) matching in the IFN group to obtain a more reliable estimate of the confounding effect of glucocorticoids on IFN beta-1a treatment. The propensity score was calculated based on a logistic regression model including age, gender, ARDS severity, Acute Physiology and Chronic Health Evaluation (APACHE) II and Sequential Organ Failure Assessment (SOFA) scores. The propensity score was included in the final logistic regression model to evaluate independent factors related to 28-day mortality. Furthermore, we present a comparison between propensity-matched patient groups. We present the associations with 28-day mortality as odds ratios (95% confidence intervals, CIs).

### Organ and cell cultures

Human lung experiments were repeated as previously presented by Bellingan et al.[9] except that hydrocortisone treatment was added as indicated below. In brief, a lung specimen from 11 different individuals were obtained by post-surgical resection of lung tissue (typically for cancer resections). The lung sections were from lung regions having normal macro- and microscopic appearance and were used with the permission from the ethics authorities at Turku University Hospital (Finland). Several small pieces were cut from all samples and 5-6 pieces/well/condition were cultured for 1 and 4 days in 24 well plates containing 1 ml of RPMI medium (supplemented with 10% fetal calf serum, 4 mM L-glutamine, 100 U/ml penicillin, and 100 μg/ml streptomycin), IFN beta (1000 UI/ml FP1201 or the drug substance (Faron Pharmaceuticals) or placebo with and without hydrocortisone (40μg/ml, Solucortef, Pfizer). FP1201 is a lyophilized form of the drug substance. After culturing, all pieces in each well were collected and frozen in OCT.

On day 1, human pulmonary endothelial cells (PromoCell) were plated 50 000 cells/well in endothelial cell growth MV medium (C-22020, PromoCell) with the supplement mix (C-19225, PromoCell) and penicillin/streptomycin to 8 well ibidi plates (Falcon). On day 2, hydrocortisone 20μg/ml (Solucortef, Pfizer) was added to half of the wells. On day 3, IFN beta (FP1201, 1000IU/ml from Faron or Rebif, 1000IU/ml from Merck) or placebo and incubation was continued for one day. Two different lots of the cells were used.

### Immunohistochemistry and immunofluorescence

Six μm thick sections were cut from cultured lung pieces and fixed with acetone. Thereafter, the sections were stained using monoclonal anti-CD73 antibody (4G4) or a negative class matched control antibody, both 10 μg/ml followed by Alexa Fluor488 conjugated goat anti-mouse IgG (Invitrogen) or HRP conjugated anti-mouse IgG (DAKO). Diaminobenzidine was used as a chromogen in peroxidase staining. The number of CD73 positive vessels was counted using stained sections and on average 20 fields/sample were counted with 200x magnification using a fluorescence microscope (Olympus).

Human pulmonary cells were washed with phosphate buffered saline (PBS) and fixed with 4% paraformaldehyde followed by treatments with 0,1 M glycine in PBS. Thereafter, the cells were incubated for 15 min in PBS containing 0.1% Triton-X and stained using anti-IRF9 antibody from LS Bio (LS-C155416) or rabbit Ig as a negative control. The second stage antibody was FITC-anti-rabbit IgG (Sigma). The signal was intensified by using Alexa Fluor 488-conjugated anti-FITC. The samples were mounted with Mowiol containing 2.5% DABCO (Sigma).

### Gel electrophoresis and immunoblotting

The nuclear and cytoplasmic fractions of human pulmonary endothelial cells with or without IFN beta-1a (1000IU/ml) ± hydrocortisone (20μg/ml) were isolated using 0.1% Triton X-100 and centrifugation at 11000 rpm. Hydrocortisone was added one day before IFN beta-1a and the pulmonary endothelial cells were lysed two hours after addition of IFN beta-1a. The protein concentration of all samples was determined by BCA protein assay kit (Pierce) and the same amount of protein was loaded on to the gel (SDS-Page). Blotting was performed using BioRad blotting system (Trans-Blot Turbo Transfer System). Blocking of the non-specific binding of the nitrocellulose transfer membranes (0.2μm) was performed with 5% bovine serum albumin and 0.1% Tween 20 in Tris buffered saline. The membranes were blotted with anti-IRF9 (LS Bio), followed by HRP-conjugated anti-rabbit IgG (DAKO). GAPDH as a loading control for cytoplasmic proteins was detected with anti-GAPDH antibody (Hytest Ltd) followed by IRDye800CW donkey anti-mouse IgG (Licor) and Histone H3 as a loading control for nuclear proteins with anti-Histone H3 antibody (Cell signalling) followed by IRDye680RD goat anti-rabbit IgG (Licor). Millipore Immobilon Western was used for signal detection.

### qPCR

Human pulmonary endothelial cells treated as described for gel analyses were collected and stored in RNAprotect Cell Reagent (QIAGEN, Hilden, Germany) at -70°C. This was followed by RNA extraction with the NucleoSpin RNA kit (Macherey-Nagel, Dueren, Germany) according to the manufacturer’s protocol. For qPCR assays, the conversion of RNA to cDNA was done with SuperScript VILO cDNA Synthesis Kit (Thermo Fisher Scientific, Espoo, Finland), followed by qPCR using the TaqMan Gene Expression Assays (Thermo Fisher Scientific, Espoo, Finland) for MX1 (Hs00895608_m1), IRF9 (Hs00196051_m1), STAT1 (Hs01013996_m1), IFNAR1 (Hs01066116_m1), IFNAR2 (Hs01022059_m1), and GAPDH (Hs02758991_g1; control gene). The reactions were run using the Applied Biosystems’ Quant Studio 3 Real-Time PCR System (Thermo Fisher Scientific). The target mRNA levels were normalised to GAPDH and a fold change of relative expression from the appropriate unstimulated control was calculated using the Applied Biosytems® analysis modules in Thermo Fisher Cloud computing platform (Thermo Fisher Scientific, Espoo, Finland).

## Results

### Clinical data

Seventy-eight out of 144 patients (54%) included in the IFN beta-1a treatment arm of the INTEREST trial received glucocorticoids during the 28-day study period, 56% (44/78) at randomization (D0), 27% (21/78) during the treatment (D1–6), and 17% (13/78) after the treatment (D7 onwards). The reported reasons for administering systemic glucocorticoids were: shock 33%, anti-inflammatory 17%, ARDS 14% and other 36% (fibrosis, immunosuppression post-transplant, adrenal insuffiency, pulmonary obstruction). The most commonly used glucocorticoids were hydrocortisone 200mg per day (59%) or methylprednisolone 40 – 120mg per day (39%) when used together with IFN beta-1a. Only 1 patient received dexamethasone with IFN beta-1a and that because of lymphoma. D28 mortality for patients receiving glucocorticoids with IFN beta-1a was 39.7% compared to 10.6% for patients receiving IFN beta-1a alone (Table 1). The Kaplan-Meier curves of the IFN beta-1a treatment arm adjusted by ARDS severity and divided according to the overlapping (D0 – D6) use of glucocorticoids with IFN beta-1a treatment demonstrate significantly increased mortality by glucocorticoid use (p = 0.0002, Figure 1). Patients who received glucocorticoids were more severely ill according to SOFA and APACHE II scores, but no significant difference in vasopressor use, P/F ratio, or in ventilation settings (Table 1). The demographic data on propensity-matched patients (glucocorticoids vs. no glucocorticoids) are presented in Table 2.

**Table 1.** Baseline demographics and 28-day mortality in the IFN beta-1a arm of the INTEREST Study divided by use of glucocorticoids during IFN treatment (D0 – D6)

| **Demographics** | **IFN beta-1 (N=144)** |  |
| --- | --- | --- |
| Concomitant glucocorticoid use with IFN beta-1a (D0-D6) | No (n=79) | Yes (n=65) |
| Age – years | 57 (18) | 59 (17) |
| Male sex - no. (%) | 55 (70) | 47 (72) |
| Female sex - no. (%) | 24 (30) | 18 (28) |
| Predisposing diagnosis for ARDS – no. (%) |  |  |
| Pneumonia | 48 (61) | 43 (66) |
| Sepsis | 9 (11) | 15 (23) |
| Aspiration | 12 (15) | 3 (5) |
| Trauma/Burns (< 15%) | 4 (5) | 0 (0) |
| Acute pancreatitis | 4 (5) | 2 (3) |
| Multiple transfusion | 2 (3) | 1 (2) |
| Other | 0 (0) | 1 (2) |
| APACHE II score – median (IQR) | 20 (8) | 24 (9) |
| SOFA score – median (IQR) | 8 (4) | 10 (6) |
| Support at randomization – no. (%) |  |  |
| Vasopressor support | 56 (71) | 57 (88) |
| Renal replacement therapy | 2 (3) | 8 (12) |
| Prone positioning | 19 (24) | 11 (17) |
| Neuromuscular blocking agents | 24 (30) | 18 (28) |
| Severe ARDS – no. (%) | 13 (16) | 15 (23) |
| Moderate ARDS – no. (%) | 66 (84) | 50 (77) |
| Ventilator settings before the first study drug dose |  |  |
| Tidal volume – ml | 443 (115) | 417 (79) |
| Positive end-expiratory pressure – cm of water | 10.5 (3.4) | 10.7 (3.6) |
| Inspiratory pressure – cm of water | 28.9 (6.9) | 31.0 (6.5) |
| Day 28 mortality | 10.6% | 39.7% |

**Table 2.** Baseline demographics and 28-day mortality for propensity-matched IFN beta-1a treated patients with and without glucocorticoids in the INTEREST Study

| Characteristics | Matched dataset (n=98) |  | Difference** |
| --- | --- | --- | --- |
|  | Concomitant glucocorticoids (n=49) | No concomitant glucocorticoids (n=49) |  |
| Age – years | 58.4 (17.4) | 58.4 (18.8) | 0.0 (18.1) |
| Male sex - no. (%) | 33 (67) | 37 (76) | -9% |
| Female sex - no. (%) | 16 (33) | 12 (24) | +9% |
| APACHE II score, range | 21.4 (6.4), 9-35 | 21.0 (6.8), 4-40 | 0.3 (6.6) |
| SOFA score, range | 9.9 (3.3), 4-20 | 9.2 (3.4), 3-19 | 0.7 (3.3) |
| Propensity score | 0.34 (0.11) | 0.33 (0.11) | 0.0 (0.11) |
| Predisposing diagnosis for ARDS – no. (%) |  |  |  |
| Pneumonia | 34 (69) | 32 (65) | 4% |
| Sepsis | 10(20) | 8 (16) | 4% |
| Aspiration | 2 (4) | 6 (12) | -8% |
| Trauma/Burns (<15%) | 0 (0) | 1 (2) | -2% |
| Acute pancreatitis | 2 (4) | 1 (2) | 2% |
| Multiple transfusion | 0 (0) | 1 (2) | -2% |
| Other*** | 1 (2) | 0 (0) | 2% |
| Severe ARDS – no. (%) | 13 (27) | 7 (14) | 13% |
| Moderate ARDS – no. (%) | 36 (73) | 42 (86) | -13% |
| 28-day mortality – no (%) | 20 (41) | 7 (14) | 27% |

\* Continuous values are presented as mean (SD) unless otherwise indicated.
\*\* T-test for continuous and Chi2-test for categorical variables. All non-significant, except for mortality (p = 0.004).

**Figure 1.**
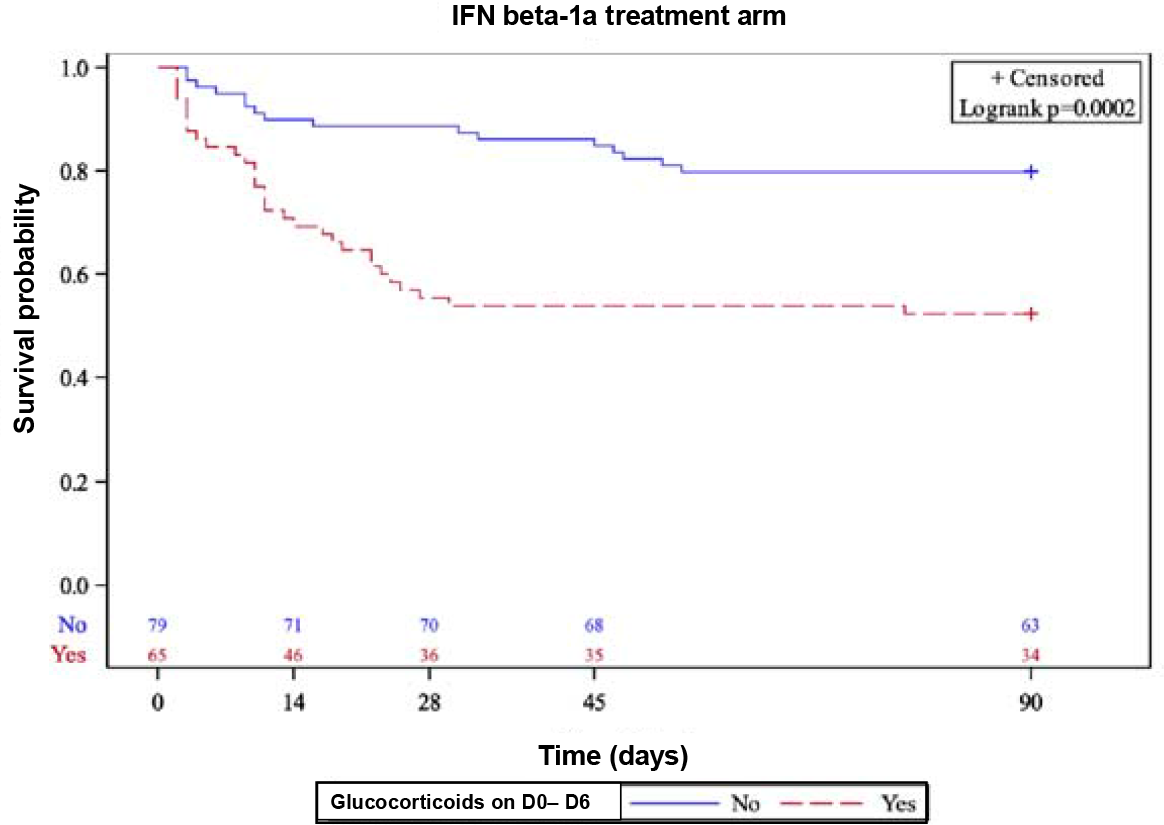
Kaplan-Meier curves of the INTEREST trial IFN beta-1a treatment arm adjusted by ARDS severity and divided according to the use overlapping (D0 – D6) use of glucocorticoids with IFN beta-1a treatment.

In the post-hoc propensity-matched analysis of the IFN beta-1a arm (n=144), baseline systemic glucocorticoid treatment was independently associated with D28 mortality (OR 5.4, 95% CI 2.1 – 13.9, P < 0.001) according to logistic regression. When the propensity matched analysis was performed using an exact matching (a precision of 0.01 propensity units), there were 49 pairs of patients who received or did not receive glucocorticoids (n=98). Among these patients, OR for increased mortality was 4.6 (95% CI 1.6 – 13.5) for those who had baseline systemic glucocorticoid treatment and 3.5 (95% CI 1.0 – 12.0) for those who initiated glucocorticoid treatment while receiving IFN beta-1a (Table 3). Glucocorticoid treatment after IFN beta-1a treatment (D7 onwards) was not associated with increased mortality.

**Table 3.**
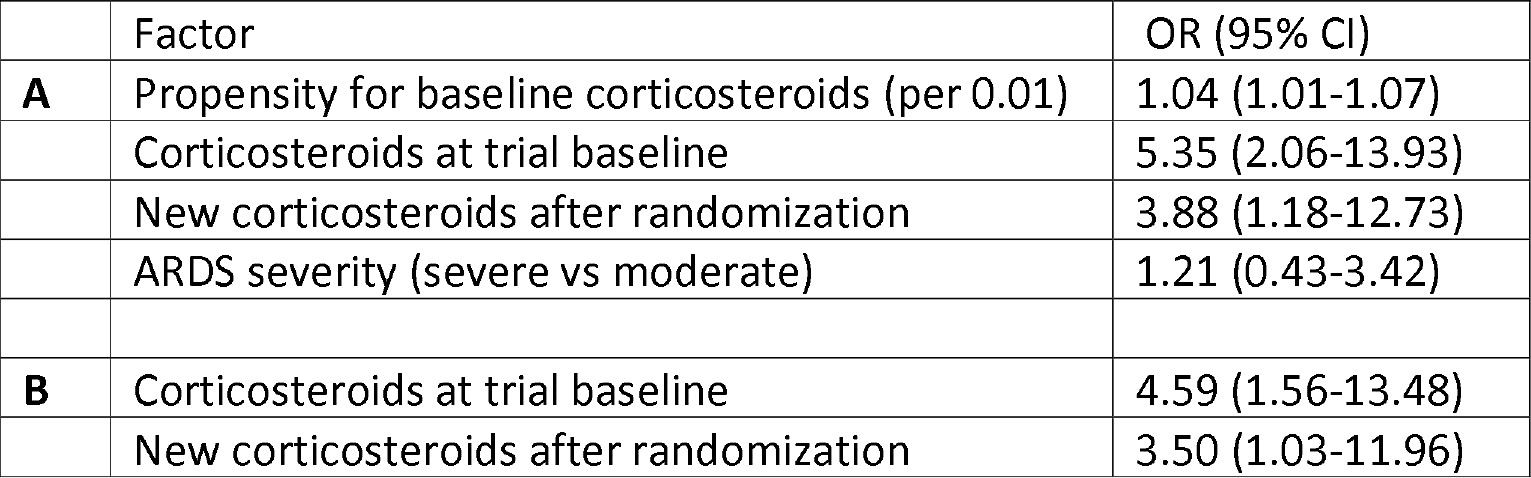
*(A)* Independent factors associated with D28 mortality in all IFN beta-1a treated patients (n=144) by logistic regression adjusted for propensity score and ARDS severity (odds ratios, OR with 95% confidence intervals, CI). *(B)* Association of glucocorticoid treatment with D28 mortality in only propensity-matched IFN beta-1a treated patients (n=98) by logistic regression (OR with 95%, CI)

|  | Factor | OR (95% CI) |
| --- | --- | --- |
| **A** | Propensity for baseline corticosteroids (per 0.01) | 1.04 (1.01-1.07) |
|  | Corticosteroids at trial baseline | 5.35 (2.06-13.93) |
|  | New corticosteroids after randomization | 3.88 (1.18-12.73) |
|  | ARDS severity (severe vs moderate) | 1.21 (0.43-3.42) |
| **B** | Corticosteroids at trial baseline | 4.59 (1.56-13.48) |
|  | New corticosteroids after randomization | 3.50 (1.03-11.96) |

### Hydrocortisone inhibits CD73 upregulation in lung organ cultures

We tested the possible inhibitory effects of hydrocortisone (HC) on IFN beta-1a induced upregulation of CD73 on blood vessels by culturing histologically normal lung tissue in the presence of IFN beta-1a with or without HC over a 4 day time course. Two different formulations of IFN beta-1a were used, FP-1201, a lyophilized product used in the phase III trial and the drug substance (DS) before lyophilization. When IFN beta-1a was applied to the culture medium CD73 expression was upregulated in comparison to the cultures without IFN beta-1a, and both pharmaceutical forms of IFN beta-1a induced a similar level of CD73 upregulation. However, in the presence of hydrocortisone, CD73 upregulation was inhibited (Figure 2 a,b).

**Figure 2.**
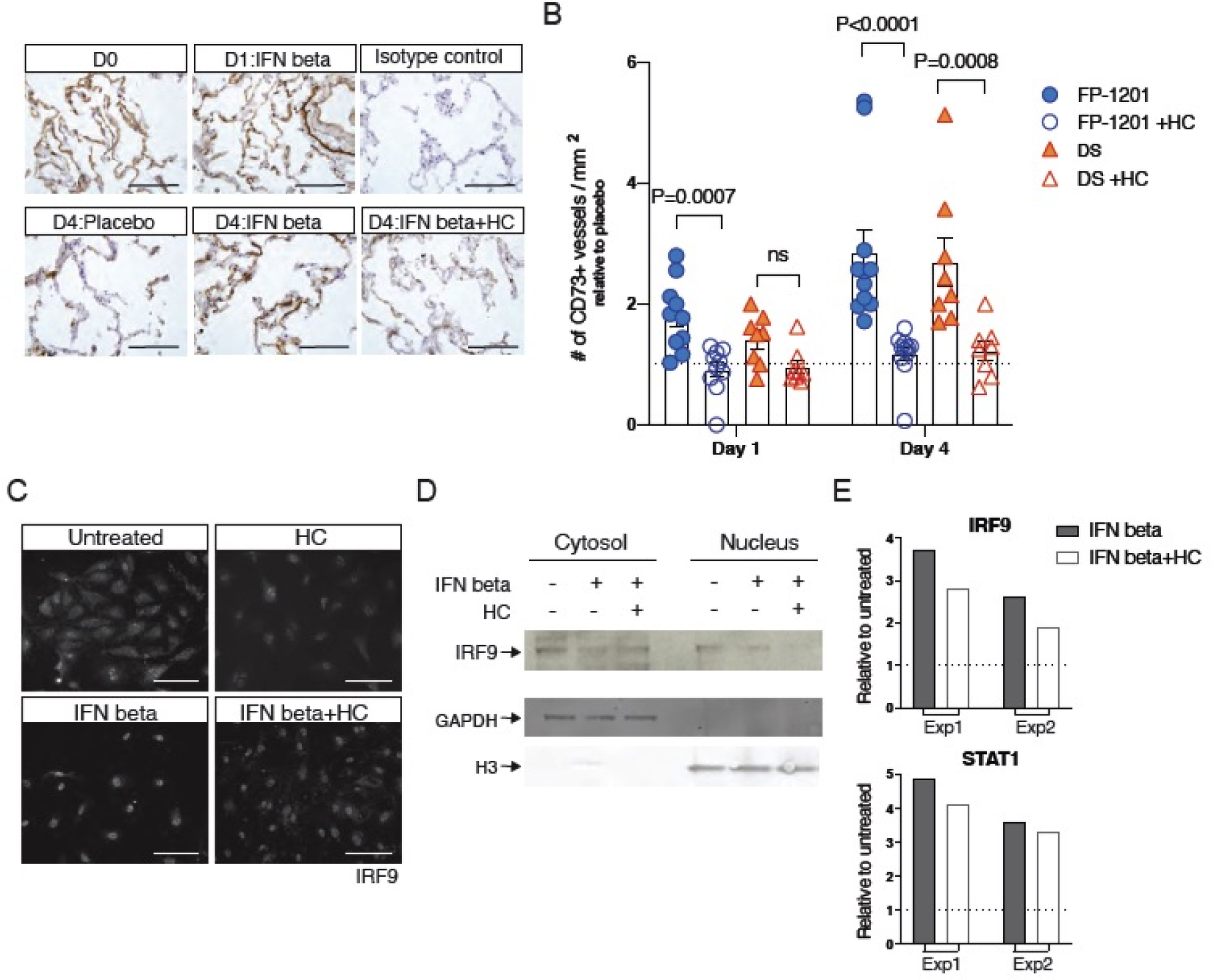
Hydrocortisone (HC) blunts IFN beta induced CD73 upregulation in lung vasculature and signaling pathways in pulmonary endothelial cells. **a** Immunohistochemical staining of CD73 (brown) in lung tissue incubated in the presence of IFN beta with or without HC for 1 and 4 days (D). D0 indicates baseline expression of CD73 in fresh lung tissue. **b** Quantification of CD73 positive vessels/mm^2^. The samples were incubated in the presence of two different IFN beta-1a formulations (FP-1201, circles n=10 (day 1) and n=11 (day 4); DS, triangles n=8). Each data point marks an individual. The dotted line represent the placebo induced level of CD73 expression. Statistical significance was analyzed using Mann Whitney test. Scale bars 100μm. **c** Immunofluorescence staining of IRF9 in primary human pulmonary endothelial cells after IFN beta treatment with or without HC showing marked inhibition of nuclear translocation of IRF9 by HC, representative results of three independent ones with similar results. **d** Immunoblotting of cytosolic and nuclear fractions of pulmonary endothelial cells after IFN beta-1a ± HC demonstrating both decrease in the signal of IRF9 and its translocation. Specific IRF9 band is indicated by an arrow, A representative blot of five independent analyses with comparable results. GAPDH and H3 (histone) are loading controls for the cytoplasmic and nuclear fractions, respectively. **e** qPCR results of two independent experiments demonstrating the decrease of IFN beta-1a triggered mRNA synthesis of IRF9 and STAT1 by HC.

### Hydrocortisone blocks IRF9 nuclear translocation and consequently IFN beta dependent signaling pathways in pulmonary lung endothelial cells

IFN beta-1a signaling via its receptor leads to the formation of a heterotrimeric transcription complex ISGF3 containing STAT1-STAT2 and IRF9, which then enters the nucleus and binds to the IFN beta responsive elements of several genes [11] or assembly on DNA [12]. As the main target of glucocorticoids is the type I interferon pathway in pulmonary epithelium [13], we tested whether this is also true in the endothelium. Primary human pulmonary endothelial cells were cultured in the presence of IFN beta-1a with and without HC or culture medium only as a baseline control. The expression of IRF9 was analyzed by immunofluorescence staining. As expected, IFN beta-1a induced the translocation of IRF9 into the nucleus, which was reduced by HC treatment (Fig 2c). To further confirm these findings, we isolated the nuclear and cytoplasmic compartments of the pulmonary endothelial cells and analyzed them separately for IRF9 by SDS-PAGE and subsequent immunoblotting (Fig 2d). Similarly, also these analyses show IRF9 translocation into the nucleus subsequent to IFN beta-1a treatment, which could be reduced by HC. In addition, HC treatment decreased IRF9 and STAT1 mRNA synthesis when measured by qPCR (Fig 2e), which was also seen at the protein level in Fig. 2c. In contrast, HC did not decrease the mRNA synthesis of the IFN alfa/beta receptor subunits as the mRNA synthesis of the IFN alfa/beta receptor (IFNAR1 and IFNAR2 subunits) increased 9 and 20%, respectively, subsequent to HC addition.

## Discussion

Our clinical and experimental data suggest an association between glucocorticoid treatment and increased D28 mortality that might be related to the hydrocortisone-induced inhibition of CD73 upregulation in human lung tissue and IFN beta dependent signaling pathways in pulmonary endothelial cells.

The use of steroids in patients with ARDS has generated great controversies [14]. While early studies suggested a clinical benefit [15] subsequent studies did not support the routine use of steroids for early [16] and persistent [17] ARDS. Our findings are consistent with a contemporary Bayesean meta-analysis that showed steroids are ineffective/harmful for ARDS [18] but contradict the results from a recent RCT on dexamethasone in ARDS patients [19]. Notable is that the dexamethasone study by Villar et al. excluded patients that were already receiving steroids at enrollment. Other possible explanations for all these inconsistent findings may lay on the origin of ARDS (viral vs. other) [20-23], genetic factors, or the use of different corticosteroids. The use of IFN beta treatment in INTEREST trial may shed light to this complex issue as only hydrocortisone and methylprednisolone were used and the majority of patients were already receiving them prior to receiving IFN beta-1a.

Increasing extracellular adenosine by activation of CD73 on epithelial and endothelial cells is one of the biological targets of IFN beta-1a. Adenosine is one of the physiological regulators of endothelial cell permeability and accelerates alveolar fluid reabsorption and inhibits leukocyte recruitment [24]. Although a previous study has shown that steroids inhibit the effect of type I IFN signaling in asthmatic epithelium [13], the use of corticosteroids was unexpectedly high (64.5% in placebo and 54.2% in the IFN beta-1a group) in the INTEREST study. Therefore, the present analyses focused on the possible interaction between steroids and IFN beta-1a in human pulmonary endothelium and CD73 expression, which a key enzyme for local adenosine production and vascular integrity. We found a significant increase in the risk of death for the baseline systemic glucocorticoid treatment and for glucocorticoid use overlapping with the IFN beta-1a treatment (Table 3). Since type I IFNs (IFN beta and alpha) are endogenously produced and are of utmost importance to fight against infections (bacterial and viral)[4], these observations may explain the fact that glucocorticoid use for patients with virus triggered ARDS is associated with an increased risk of death [16]. In line with these interpretations, others have shown that the use of recombinant human IFN beta inhibits SARS virus replication and has antiviral potential even after infection [25]. Furthermore, the use of IFN alpha was associated with lower mortality in the previous MERS epidemic [26], but IFNs failed to repeat the results in a larger cohort of MERS patients [27]. However, these patients received sub-cutaneous IFN beta which gives limited direct drug exposure to lung endothelial cells. More importantly, 60% of these patients received corticosteroids, which based on our findings prevents IFN beta signaling.

Earlier reports shown that glucocorticoids target type I interferon signaling pathways by inhibiting the assembly of the ISGF3 complex in mouse macrophages and human epithelial cells [11, 13, 28]. In our pulmonary endothelial cell cultures, we observed that, the use of hydrocortisone for one day reduced the mRNA synthesis of IRF9 and STAT1 – the components of the ISGF3 complex. The effect was not due to the downregulation of IFN alpha/beta receptor. Moreover, we witnessed inhibition of IRF9 translocation into the nucleus, which is a requirement for triggering IFN beta responsive genes. IFR9 is essential for type I IFN response as its’ deficiency leads to severe life-threatening conditions by common viral infection [29]. As there are more than 7300 type 1 interferon responsive genes (http://www.interferome.org/interferome/site/dbStat.jspx) the effects of glucocorticoids are extensive already when taken only their targeting of type 1 IFN signaling pathways into account. In addition, glucocorticoids inhibit the production and secretion of type I IFNs themselves [30]. We used hydrocortisone in ex vivo experiments as it is commonly used in clinical practice. Although glucocorticoids have different potencies, they all act via the glucocorticoid receptor and affect the same signaling pathways [31], thus the obtained effects are expected to be generalized to all glucocorticoids used to treat ARDS patients.

As STAT1-STAT2-IRF9 complex also drives the expression of HIF-1a [32], it can be envisioned that also the expression of HIF-1a is decreased due to glucocorticoid use. In addition, HIF-1a mediated transactivation of genes may also be impacted as glucocorticoids induce the phosphorylation of GRIP1 [33], a normal interaction required for HIF-1a mediated gene transactivation. These earlier data together with our *ex vivo* results utilizing hydrocortisone together with IFN beta-1a demonstrate that glucocorticoids interfere with the production of CD73, since the gene encoding for CD73 contains response elements for interferons (ISRE) and hypoxia (HRE) [34, 35].

In conclusion, we found that hydrocortisone inhibits CD73 upregulation in human lung tissue and IFN beta dependent signaling pathways in pulmonary endothelial cells. CD73 is an important regulator of the vascular integrity especially needed in conditions such as ARDS. Therefore, these experimental findings, in combination with our propensity-matched analysis from IFN beta-1a treated patients from the INTERERST trial, provide a mechanistic evidence of the harmful effect of glucocorticoids on the IFN response in the human lung. These data provides a rationale for contraindication to use steroids in patients with ARDS of viral origin, when an IFN response is required.

## Data Availability

The data and the novel reagent (anti-CD73 antibody) will be made available to others on reasonable requests to the corresponding author

## Declarations

### Ethics approval and consent to participate

All procedures performed in studies involving human participants in the original INTEREST trial (ClincalTrials.gov Identifier NCT02622724) were in accordance with the ethical standards of the institutional and/or national research committees and the Helsinki declaration.

### Consent for publication

Not applicable

### Availability of data and material

After publication, the data and the novel reagent (anti-CD73 antibody) will be made available to others on reasonable requests to the corresponding author.

### Competing interests

JJ, MK, JM and MJ are employees and shareholders of Faron Pharmaceuticals. MH and SJ own stocks of Faron Pharmaceuticals and SJ has a patent (US 7534423).

## Funding

Academy of Finland, Faron Pharmaceuticals Ltd, European Union Seventh Framework Program (grant agreement No. 305853)

## Author contributions

KE, MH and SJ contributed to the experimental study design and analyzed the results of the experiments. JJ, VP, MK, JM,MJ, GB and VMR contributed to the original INTREST trial and to the post-hoc analyses presented in this paper. MM consented the patients donating the lung specimens and performed the surgical operations. TH performed the statistical analyses. JJ, VP and SJ wrote the first draft of the manuscript. All authors contributed to its final version.

## Acknowledgements

We thank Riikka Sjöroos and Sari Mäki for expert technical assistance and the study group of the INTEREST trial for participating to the study at the trial sites.

## Abbreviations

(APACHE) II: Acute Physiology and Chronic Health Evaluation
(IFN(: interferon
(SOFA): Sequential Organ Failure Assessment

## Notes

### Clinical Trial

The original data are from INTEREST trial: ClincalTrials.gov Identifier NCT02622724

## References

1. Pham T, Rubenfeld GD, (2017) Fifty Years of Research in ARDS. The Epidemiology of Acute Respiratory Distress Syndrome. A 50th Birthday Review. Am J Respir Crit Care Med 195: 860–870

2. Matthay MA, McAuley DF, Ware LB, (2017) Clinical trials in acute respiratory distress syndrome: challenges and opportunities. Lancet Respir Med 5: 524–53438.

3. Huppert LA, Matthay MA, Ware LB, (2019) Pathogenesis of Acute Respiratory Distress Syndrome. Semin Respir Crit

4. Singanayagam A, Glanville N, Girkin JL, Ching YM, Marcellini A, Porter JD, Toussaint M, Walton RP, Finney LJ, Aniscenko J, Zhu J, Trujillo-Torralbo MB, Calderazzo MA, Grainge C, Loo SL, Veerati PC, Pathinayake PS, Nichol KS, Reid AT, James PL, Solari R, Wark PAB, Knight DA, Moffatt MF, Cookson WO, Edwards MR, Mallia P, Bartlett NW, Johnston SL, (2018) Corticosteroid suppression of antiviral immunity increases bacterial loads and mucus production in COPD exacerbations. Nat Commun 9: 2229

5. Salmi M, Jalkanen S, (2014) Ectoenzymes in leukocyte migration and their therapeutic potential. Semin Immunopathol 36: 163–176

6. Niemela J, Ifergan I, Yegutkin GG, Jalkanen S, Prat A, Airas L, (2008) IFN-beta regulates CD73 and adenosine expression at the blood-Care Med 40: 31-39brain barrier. Eur J Immunol 38: 2718–2726

7. Kiss J, Yegutkin GG, Koskinen K, Savunen T, Jalkanen S, Salmi M, (2007) IFN-beta protects from vascular leakage via up-regulation of CD73. Eur J Immunol 37: 3334–3338

8. Antonioli L, Pacher P, Vizi ES, Hasko G, (2013) CD39 and CD73 in immunity and inflammation. Trends in Mol Med 19: 355-367

9. Bellingan G, Maksimow M, Howell DC, Stotz M, Beale R, Beatty M, Walsh T, Binning A, Davidson A, Kuper M, Shah S, Cooper J, Waris M, Yegutkin GG, Jalkanen J, Salmi M, Piippo I, Jalkanen M, Montgomery H, Jalkanen S, (2014) The effect of intravenous interferon-beta-1a (FP-1201) on lung CD73 expression and on acute respiratory distress syndrome mortality: an open-label study. Lancet Respir Med 2: 98-107

10. Ranieri VM, Pettilä V, Karvonen MK, Jalkanen J, Nightingale P, Brealey D, Mancebo J, Ferrer R, Mercat A, Patroniti N, Quintel M, Vincent JL, Okkonen M, Meziani F, Bellani G, MacCallum N, Creteur J, Kluge S, Artigas-Raventos A, Maksimow M, Piippo I, Elima K, Jalkanen S, Jalkanen M, Bellingan G, Group IS, (2020) Effect of Intravenous Interferon β-1a on Death and Days Free From Mechanical Ventilation Among Patients With Moderate to Severe Acute Respiratory Distress Syndrome: A Randomized Clinical Trial. JAMA doi: 10.1001/jama.2019.22525 (ePub ahead of print)

11. Flammer JR, Dobrovolna J, Kennedy MA, Chinenov Y, Glass CK, Ivashkiv LB, Rogatsky I, (2010) The type I interferon signaling pathway is a target for glucocorticoid inhibition. Mol Cell Biol 30: 4564–4574

12. Platanitis E, Demiroz D, Schneller A, Fischer K, Capelle C, Hartl M, Gossenreiter T, Müller M, Novatchkova M, Decker T, (2019) A molecular switch from STAT2-IRF9 to ISGF3 underlies interferon-induced gene transcription. Nat Commun 10: 2921

13. Diez D, Goto S, Fahy JV, Erle DJ, Woodruff PG, Wheelock Å, Wheelock CE, (2012) Network analysis identifies a putative role for the PPAR and type 1 interferon pathways in glucocorticoid actions in asthmatics. BMC Med Genomics 5: 27

14. Thompson BT, Ranieri VM, (2016) Steroids are part of rescue therapy in ARDS patients with refractory hypoxemia: no. Intensive Care Med 42: 921–923

15. Meduri GU, Headley AS, Golden E, Carson SJ, Umberger RA, Kelso T, Tolley EA, (1998) Effect of prolonged methylprednisolone therapy in unresolving acute respiratory distress syndrome: a randomized controlled trial. JAMA 280: 159–165

16. Ruan SY, Lin HH, Huang CT, Kuo PH, Wu HD, Yu CJ, (2014) Exploring the heterogeneity of effects of corticosteroids on acute respiratory distress syndrome: a systematic review and meta-analysis. Crit Care 18: R63

17. Steinberg KP, Hudson LD, Goodman RB, Hough CL, Lanken PN, Hyzy R, Thompson BT, Ancukiewicz M, National Heart Ln, and Blood Institute Acute Respiratory Distress Syndrome (ARDS) Clinical Trials Network, (2006) Efficacy and safety of corticosteroids for persistent acute respiratory distress syndrome. N Engl J Med 354: 1671–1684

18. Peter JV, John P, Graham PL, Moran JL, George IA, Bersten A, (2008) Corticosteroids in the prevention and treatment of acute respiratory distress syndrome (ARDS) in adults: metaanalysis. BMJ 336: 1006–1009

19. Villar J, Ferrando C, Martínez D, Ambrós A, Muñoz T, Soler JA, Aguilar G, Alba F, González-Higueras E, Conesa LA, Martín-Rodríguez C, Díaz-Domínguez FJ, Serna-Grande P, Rivas R, Ferreres J, Belda J, Capilla L, Tallet A, Añón JM, Fernández RL, González-Martín JM, network diA, (2020) Dexamethasone treatment for the acute respiratory distress syndrome: a multicentre, randomised controlled trial. Lancet Respir Med 8: 267–276

20. Brun-Buisson C, Richard JC, Mercat A, Thiébaut AC, Brochard L, Group R-SAHNvR, (2011) Early corticosteroids in severe influenza A/H1N1 pneumonia and acute respiratory distress syndrome. Am J Respir Crit Care Med 183: 1200–1206

21. Takaki M, Ichikado K, Kawamura K, Gushima Y, Suga M, (2017) The negative effect of initial high-dose methylprednisolone and tapering regimen for acute respiratory distress syndrome: a retrospective propensity matched cohort study. Crit Care 21: 135

22. Zhang Z, Chen L, Ni H, (2015) The effectiveness of Corticosteroids on mortality in patients with acute respiratory distress syndrome or acute lung injury: a secondary analysis. Sci Rep 5: 17654

23. Moreno G, Rodríguez A, Reyes LF, Gomez J, Sole-Violan J, Díaz E, Bodí M, Trefler S, Guardiola J, Yébenes JC, Soriano A, Garnacho-Montero J, Socias L, Del Valle Ortíz M, Correig E, Marín-Corral J, Vallverdú-Vidal M, Restrepo MI, Torres A, Martín-Loeches I, Group GS, (2018) Corticosteroid treatment in critically ill patients with severe influenza pneumonia: a propensity score matching study. Intensive Care Med 44: 1470–1482

24. Eltzschig HK, (2013) Extracellular adenosine signaling in molecular medicine. J Mol Med (Berl) 91: 141–146

25. Cinatl J, Morgenstern B, Bauer G, Chandra P, Rabenau H, Doerr HW, (2003) Treatment of SARS with human interferons. Lancet 362: 293–294

26. Omrani AS, Saad MM, Baig K, Bahloul A, Abdul-Matin M, Alaidaroos AY, Almakhlafi GA, Albarrak MM, Memish ZA, Albarrak AM, (2014) Ribavirin and interferon alfa-2a for severe Middle East respiratory syndrome coronavirus infection: a retrospective cohort study. Lancet Infect Dis 14: 1090–1095

27. Arabi YM, Shalhoub S, Mandourah Y, Al-Hameed F, Al-Omari A, Al Qasim E, Jose J, Alraddadi B, Almotairi A, Al Khatib K, Abdulmomen A, Qushmaq I, Sindi AA, Mady A, Solaiman O, Al-Raddadi R, Maghrabi K, Ragab A, Al Mekhlafi GA, Balkhy HH, Al Harthy A, Kharaba A, Gramish JA, Al-Aithan AM, Al-Dawood A, Merson L, Hayden FG, Fowler R, (2019) Ribavirin and Interferon Therapy for Critically Ill Patients With Middle East Respiratory Syndrome: A Multicenter Observational Study. Clin Infect Dis doi: 10.1093/cid/ciz544. [Epub ahead of print]

28. Ogawa S, Lozach J, Benner C, Pascual G, Tangirala RK, Westin S, Hoffmann A, Subramaniam S, David M, Rosenfeld MG, Glass CK, (2005) Molecular determinants of crosstalk between nuclear receptors and toll-like receptors. Cell 122: 707–721

29. Hernandez N, Melki I, Jing H, Habib T, Huang SSY, Danielson J, Kula T, Drutman S, Belkaya S, Rattina V, Lorenzo-Diaz L, Boulai A, Rose Y, Kitabayashi N, Rodero MP, Dumaine C, Blanche S, Lebras MN, Leung MC, Mathew LS, Boisson B, Zhang SY, Boisson-Dupuis S, Giliani S, Chaussabel D, Notarangelo LD, Elledge SJ, Ciancanelli MJ, Abel L, Zhang Q, Marr N, Crow YJ, Su HC, Casanova JL, (2018) Life-threatening influenza pneumonitis in a child with inherited IRF9 deficiency. J Exp Med 215: 2567–2585

30. Bhattacharyya S, Zhao Y, Kay TW, Muglia LJ, (2011) Glucocorticoids target suppressor of cytokine signaling 1 (SOCS1) and type 1 interferons to regulate Toll-like receptor-induced STAT1 activation. Proc Natl Acad Sci U S A 108: 9554–9559

31. Nicolaides NC, Pavlaki AN, Maria Alexandra MA, Chrousos G. (2018) Glucocorticoid Therapy and Adrenal Suppression. In: Endotext [Internet]. South Dartmouth (MA): MDText.com, Inc 2000-2018 PMID: 25905379

32. Gerber SA, Pober JS, (2008) IFN-alpha induces transcription of hypoxia-inducible factor-1alpha to inhibit proliferation of human endothelial cells. J Immunol 181: 1052–1062

33. Dobrovolna J, Chinenov Y, Kennedy MA, Liu B, Rogatsky I, (2012) Glucocorticoid-dependent phosphorylation of the transcriptional coregulator GRIP1. Mol Cell Biol 32: 730–739

34. Synnestvedt K, Furuta GT, Comerford KM, Louis N, Karhausen J, Eltzschig HK, Hansen KR, Thompson LF, Colgan SP, (2002) Ecto-5’-nucleotidase (CD73) regulation by hypoxia-inducible factor-1 mediates permeability changes in intestinal epithelia. J Clin Invest 110: 993–1002

35. Niemela J, Henttinen T, Yegutkin GG, Airas L, Kujari AM, Rajala P, Jalkanen S, (2004) IFN-alpha induced adenosine production on the endothelium: a mechanism mediated by CD73 (ecto-5’-nucleotidase) up-regulation. J Immunol 172: 1646–1653

